# Interventions to support parents, families and caregivers in caring for preterm or low birth weight infants at home: a systematic review

**DOI:** 10.1101/2022.10.25.22281452

**Authors:** C Bedwell, V Actis Danna, N Tate, K Dwan, A Portela, T Lavender

## Abstract

**Background:** Preterm and low birth weight (LBW) (<2500g) infants are at greater risk of mortality and morbidity. Interventions to support parents to care for their newborn infant in the home may help to improve outcomes.

**Objective:** To determine what interventions, approaches, or strategies to support mothers/fathers/caregivers and families in caring for preterm or LBW infants in the home have been effective in improving outcomes.

**Methods:** A comprehensive search of relevant electronic databases, including MEDLINE, Embase, CINAHL and Cochrane Central Register of Controlled Trials was completed in September 22. Two reviewers screened papers in Covidence and extracted data from 41 included papers. Quality of papers and certainty of evidence were assessed using CASP and GRADE, respectively.

**Results:** There is some evidence that support interventions may improve outcomes related to infant mortality, improvements in infant growth, exclusive breastfeeding, infant cognitive development, immunisation uptake, and reduction in maternal stress and depression. However, the overall certainty of evidence is low or very low in the majority of studies.

**Conclusions:** Interventions providing support for parents to care for infants in the home may improve outcomes for this population. There is a need for well-considered large scale support interventions, prioritised and developed with women and families.

## Introduction

In 2019, 2.4 million children died within 28 days of birth with a further estimated 1.5 million deaths in the first year of life (1). Infants who are born prematurely (below 37 weeks gestation) or are of low birth weight (LBW) (<2500g) are at greater risk of mortality and morbidity (2). Of those that do survive, associated long term morbidity can impact on their wellbeing decades later (3). Many advances in neonatal care have led to a reduction in infant mortality. The mortality rate for neonates has reduced from 31 per 1000 live births in 2000 to 18 per 1000 live births in 2018, but this is still short of the Sustainable Development Goal (SDG) 3 target of 12 per 1000 live births by 2030, and the global aim to end preventable deaths (1). Whilst the majority of newborns (75%, 2.4 m) die in the first week of life (1), a considerable number (1.4m) die during infancy; between 28 days to one year of age, many after facility discharge. Hence, effective interventions to improve infant health and wellbeing in the short and longer term are important in reducing mortality and morbidity.

Parents are the main carers for infants following their discharge from healthcare facilities, yet often feel ill equipped for the task of caring for a preterm or LBW infant at home (4, 5). This can be due to a lack of support, confidence, education or practical and environmental factors (6). This, in turn, can have an impact on parental wellbeing and their subsequent ability to care for their newborn. Evidence suggests that in addition to the stress of caring for a preterm or LBW infant, women are up to 40% more likely to suffer from depression than mothers of healthy infants (7). Furthermore, in some settings, stigma and community prejudice towards LBW babies may also impact on women’s ability to care for their infant without support (8). Many reported interventions are conducted within the facility setting, however, the period of time after discharge from health system care may be the most vital in terms of parental needs. It is at this point that parents report feeling overwhelmed and in need of greater support (9). Psychosocial, emotional and financial support, along with greater understanding of developmental expectations, have been raised as concerns by parents following discharge (6).

Interventions to support parents to care for their newborn may help to improve outcomes. A systematic review focussed on healthy term infants suggested that nurturing parenting interventions can improve developmental outcomes (10). A further systematic review demonstrated parent-to-parent support in facility neonatal care increased perceptions of support, reduced maternal stress, and increased mothers’ confidence in the ability to care for their baby, but had no impact on parenting practices nor on breastfeeding (11). This review will focus specifically on interventions that aim to support parents to care for their newborn preterm or LBW infant in the home.

## Methods

The systematic review followed standard systematic review principles and is reported in line with PRISMA reporting requirements (12). The review protocol is published in the International Prospective Register of Systematic Reviews (https://www.crd.york.ac.uk/PROSPEROCRD42021275525). The review was expanded from interventions taking place in the home only, to those which were initiated in the facility or in the home, but which related to outcomes measured following discharge. This was to capture effects of facility-based interventions that impacted on parental support following discharge. See supplementary file 1 for search strategy.

### Eligibility criteria

Studies were included if they met the following inclusion criteria:

- Participants were mother/father/parents, families or caregivers of preterm or LBW infants.
- Included interventions which focused on providing support to participants to care for their infants in the home; including, education and counselling, home visits, family and community mobilization approaches, policy and commenced either during pregnancy, before discharge or after discharge/after birth, with outcomes measured in the home.
- Studies which reported critical and priority infant and maternal outcomes, and which were measured in the home up to 12 months of age. Critical outcomes comprised infant mortality (number of deaths), morbidity (as defined by authors), growth (length or weight) and neurodevelopment (cognitive, motor, measured by scale). Priority outcomes comprised breastfeeding (exclusive and/or duration), care seeking (hospital visits or contact with health services), parent-infant interaction, mother-child attachment (measured by scale) and parental health and wellbeing (stress, anxiety depression, quality of life measured by scale).
- Randomised controlled trials; quasi-experimental studies with a control group; cohort studies or interrupted time series with outcome measures at a minimum of three time points.
- Studies were included regardless of high-, middle- or low-income setting.
- Were published in full text between 2000 and 2021.

Studies were excluded if their focus was on clinical care only, did not report data from preterm or LBW infants, did not have a control group or were of qualitative design, or were published before 2000, to ensure studies of current relevance were included.

A comprehensive search of relevant electronic databases included Medical Literature Analysis and Retrieval System Online (MEDLINE), Excerpta Medica Database (Embase), Cumulative Index to Nursing and Allied Health Literature (CINAHL) and Cochrane Central Register of Controlled Trials, using a detailed search strategy (see supplementary file 1). Citation searching was used to identify any further studies meeting the inclusion criteria. The search was completed between October and December 2021.

### Screening, data extraction and analysis

Search results were managed through the Covidence software platform (https://www.covidence.org/), to allow for removal of duplicates and screening. Two reviewers independently screened all papers on title and abstract, and reviewed the remaining full-text papers for inclusion, in accordance with the eligibility criteria. Any discrepancies between the reviewers were resolved during a group meeting to discuss and resolve each conflicted decision and achieve consensus.

A data extraction form, tailored to the review parameters was used in conducting data extraction, which was conducted independently and in duplicate. The data extracted was organized into domains related to the intervention type and outcome.

Data were synthesised in relation to intervention type and outcome. There was considerable heterogeneity across the studies in terms of study designs, interventions and outcomes. Studies were grouped into intervention type, based on the main characteristics of the intervention. A variety of outcome measures and timepoints were used by the included studies. For consistency, final timepoint measurements for each study were included in the analysis. Heterogeneity between studies limited the opportunity for meta-analysis, with most outcomes synthesised narratively. Where meta-analysis was possible; data were input into Revman version 5.4.1 (revman.cochrane.org). For dichotomous data, risk ratio and 95% confidence intervals were reported. For continuous data, mean difference or standardised mean difference and 95% confidence intervals were reported. Standardised mean difference was reported where different measurement scales were used. Effect size is reported in line with Cohen’s effect sizes; 0.2 small effect, 0.5 moderate effect, 0.8 large effect. Heterogeneity between studies was identified by visual inspection of forest plot, chi squared (p<0.1) and I squared. Where heterogeneity was identified, the reasons for this were explored. If there was no significant heterogeneity, we pooled the results using fixed effect model. We anticipated from the scoping review that sub-group analysis would not be possible, due to the diversity of interventions and outcomes. Non-randomised trials are reported narratively, using adjusted estimates where available.

### Risk of bias and certainty assessment

Quality assessment and risk of bias was completed using CASP (12) and the original Cochrane risk of bias tool by two reviewers, independently. Certainty of the body of evidence was assessed using Grading of Recommendations Assessment Development and Evaluation (GRADE) approach, as outlined in the GRADE Handbook (https://gdt.gradepro.org/app/handbook/handbook.html), for each reported outcome. This GRADE approach allows for evidence certainty to be downgraded according to five domains: risk of bias; inconsistency; indirectness; imprecision; and publication bias. The results of the assessment for each study relate to one of four grades: high, moderate, low or very low. Studies were included regardless of quality or certainty of evidence given the diversity of interventions, allowing for the range of studies, interventions and outcomes to be reviewed.

## Results

A total of 1752 papers were identified and screened for inclusion. After removal of duplicates and screening of title and abstract 201 papers received full-text review. A total of 41 papers, related to 37 studies, were included in the final review. See PRISMA flowchart, Figure 1 for details.

**Figure 1:**
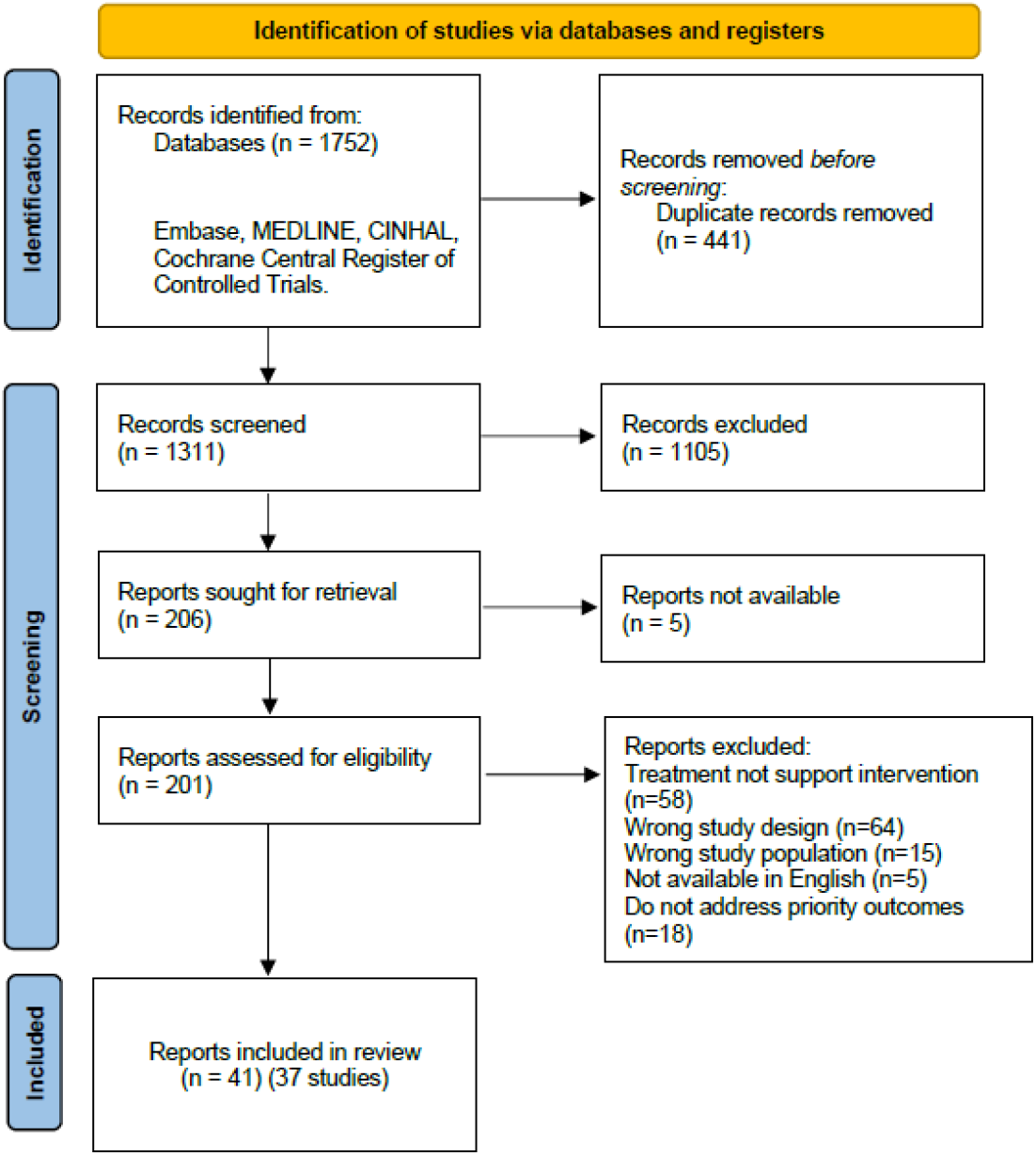
Prisma flowchart.

### Characteristics of included studies

Twenty-five studies were randomised controlled trials (RCTs), six pilot RCTs, four quasi-experimental studies and two before-and-after intervention studies, comprising a total of 11,758 participants. The majority of studies (n=27) were based in high-income settings (Australia, Canada, Denmark, Finland, Greece, Hong Kong, Netherlands, Norway, South Korea, Sweden, UK, USA), three in upper middle-income (China, Taiwan, Jamaica), and seven in lower middle-income settings (Bangladesh, Egypt, India, Iran, Philippines). None of the studies were based in low-income settings.

Risk of bias was unclear or high in many studies due to limited explanation of randomisation, allocation concealment and assessor blinding. The majority of studies were of low or very low certainty evidence, based on Grade, as a result of risk of bias, indirectness and imprecision (tables 2-6).

#### Types of interventions

The number and type of interventions were diverse, but some shared similar characteristics and were combined into groups for analysis. Five groups of interventions were identified; education and counselling interventions initiated in the health facility, home visits by a trained health worker or volunteer, discharge preparation and readiness interventions, digital communication interventions and peer support interventions. Although there was some overlap of interventions between groups, the studies were assigned to groups with which the interventions shared the greatest number of similar characteristics. All the interventions commenced either in the health facility or in the home following birth or discharge from the facility. Interventions focussed on mothers and infants, with involvement of partner and family in limited cases. Control groups provided usual care, although this varied between settings and contexts. See table 1 for a summary overview of the interventions.

##### Education and counselling interventions initiated in the health facility

Eighteen studies included in this category focused on education or training of parents to care for their newborn, either one-to-one or in groups (13–29). The intervention training topics covered aspects of responsiveness and sensitivity to the infant, understanding and promotion of infant development, care of the newborn and breastfeeding. All provided education in the facility, with nine studies continuing education in the home following discharge, with interventions provided by health workers or researchers.

##### Home visiting interventions

Nine home visiting studies comprised of interventions which commenced and continued in the home following birth (30–42). Parents were supported in aspects of their infants’ care including care planning in the home, sensitivity and responsiveness, promotion of infant development and breastfeeding. The interventions were provided by healthcare workers, community health workers, trained intervention workers or trained volunteers.

##### Strengthened discharge preparedness interventions

Four studies focused on preparing parents for the discharge of their infant, in addition to normal discharge preparation (43–46). This involved individualised discharge planning with the parents, along with advice on general care of the infant. The interventions commenced in the facility, with three continuing at home after discharge; two by telephone contact. These interventions were delivered by health workers trained in neonatal care.

##### Digital communication interventions

Four studies comprised of interventions focussing on support and communication specifically through digital means, using web-based, Skype, apps or telephone media (47–50). Parents were supported in care of the infant and breastfeeding, with interventions commencing either in the facility or at home. These were provided by health workers trained in delivery of the intervention, including neonatal intensive care unit (NICU staff), nurses and, in one study, occupational therapists.

##### Peer support interventions

Two studied comprised of interventions delivered by peer supporters who were women with experience of caring for a preterm infant in a similar environment and were willing to use their experiences to support others (51, 52). The focus was on general and breastfeeding support. These interventions commenced in the facility and took place following agreement from the mother or were initiated by the mother.

#### Outcomes

Outcomes are reported in relation to each intervention type, with accompanying evidence tables.

### Education and counselling interventions initiated in the health facility

Eighteen 18 studies were included in this intervention category. Of these, nine reported infant outcomes including infant growth (two studies) exclusive breastfeeding (two studies) cognitive development (three studies) and infant temperament (two studies) (Table 2).

An improvement in infant growth measured by length and weight was reported in two RCTs (16, 26). An increase in length of the infant (measured in cms) in the intervention group at 60 days follow up (n=184, MD 1.5 cm, 95% CI 1.1 cm to 1.9 cm) and 120 days follow up (n=57, MD 1.2 cm, 95% CI 0.2 cm to 2.6 cm) was evident. Infant weight (measured in grams) also increased in the intervention groups at 60 days (MD 305 g, 95% CI 228 g to 382 g) and 120 days (MD 410 g, 95% CI 406 g to 414 g).

An increase in exclusive breastfeeding at 2-3 months corrected age was reported by two RCTs (13, 26), favouring the intervention group (n=244, RR 1.71, 95% CI 1.26 to 2.31).

**Figure 2:**
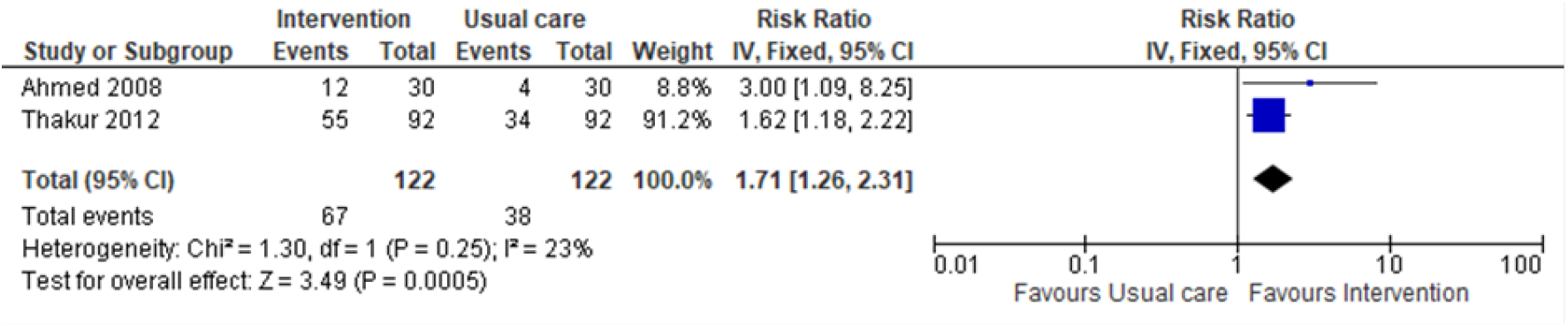
Exclusive breastfeeding at 2-3 months corrected age.

Three small RCTs (14, 17, 21) reported cognitive development at 4-6 months corrected age and were included in meta-analysis, suggesting moderate evidence of effect for those infants receiving the intervention (n=65, SMD 0.67, 95% CI 0.16 to 1.17).

**Figure 3:**
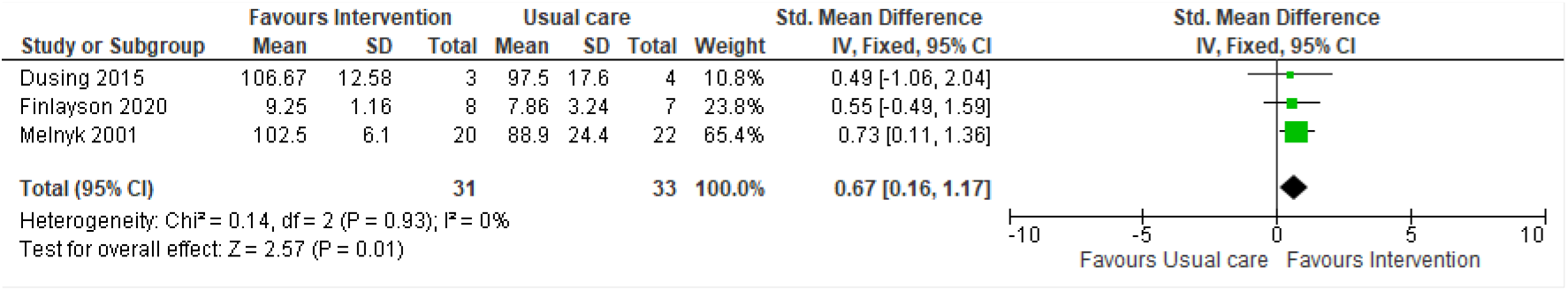
Infant cognitive development (measured by Bailey Scales of Infant Development II & III)

Two RCTs (22, 24) reported no evidence of effect in infant temperament at 6 months corrected age (n=155, MD 0.54, 95% CI −0.06 to 1.02).

Maternal outcomes were reported by 13 studies and included maternal stress (three studies) depression (three studies), anxiety (four studies) and maternal-infant interaction (four studies) (Table 2).

Three RCTs (19, 53, 54) measured stress, at differing time points, with two studies reporting no evidence of effect at 15 days (53) (n=52, MD −1.12, 95% CI −14.32 to 12.08) or three months corrected age (19) (n=199, MD 4.80, 95% CI −0.56 to 10.16). However, one study (54) reported a reduction in stress at 12 months corrected age (n=130, MD −13.70, 95% CI −25.5 to −1.89).

One RCT (53) reported no evidence of effect on maternal depression following a 15 day intervention (n=52, (MD −0.64, 95% CI −3.53 to 2.25). Two studies (21, 24) were included in meta-analysis, demonstrating small evidence of effect in reducing depression (n=94, SMD −0.25, 95% CI −0.65 to 0.16) at six months.

**Figure 3:**
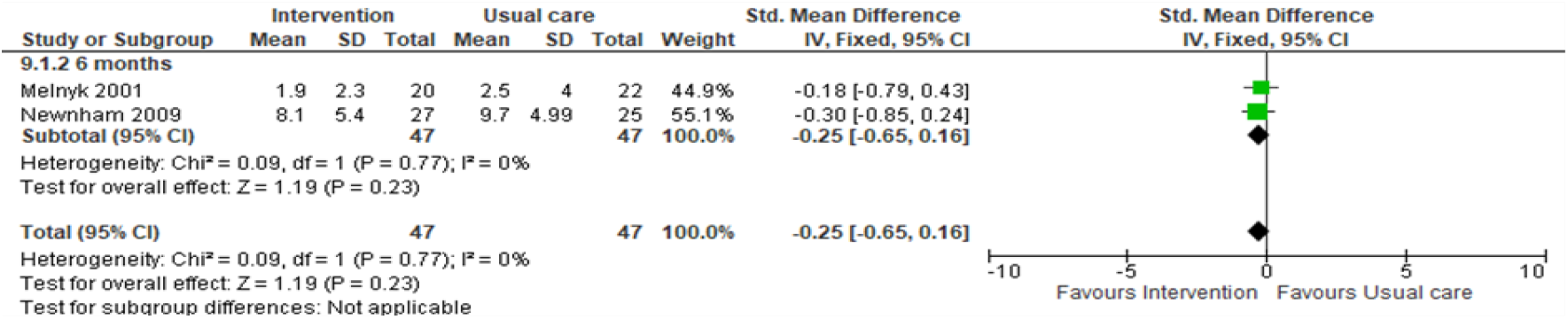
Maternal depression (measured by Edinburgh Postnatal Depression Scale and Profile of Mood States)

There was no evidence of effect reported for maternal anxiety or parent-infant interaction (Table 2).

### Home visiting interventions

Nine studies were included in this category. Of these, eight reported infant reported outcomes related to mortality (two studies), exclusive breastfeeding (three studies), cognitive (two studies) and motor development (one study) and temperament (one study) (Table 3).

One large RCT (36)(n=7479) reported a significant reduction in infant mortality in the intervention group (RR 0.71, 95% CI 0.57 to 0.89) at 180 days follow up. One observational study (39) reported a reduction in mortality over the first year of life, but this was not significant (n=970, RR 0.14 95% CI 0.02 to 1.16).

Three RCTs (30, 32, 36) included in a meta-analysis suggest no evidence of effect in exclusive breastfeeding at six months corrected age (n=7221, RR 4.48, 95% CI 0.28 to 72.9). However, two of the studies (30, 36) reported an increase in exclusive breastfeeding in the intervention group.

**Figure 4:**
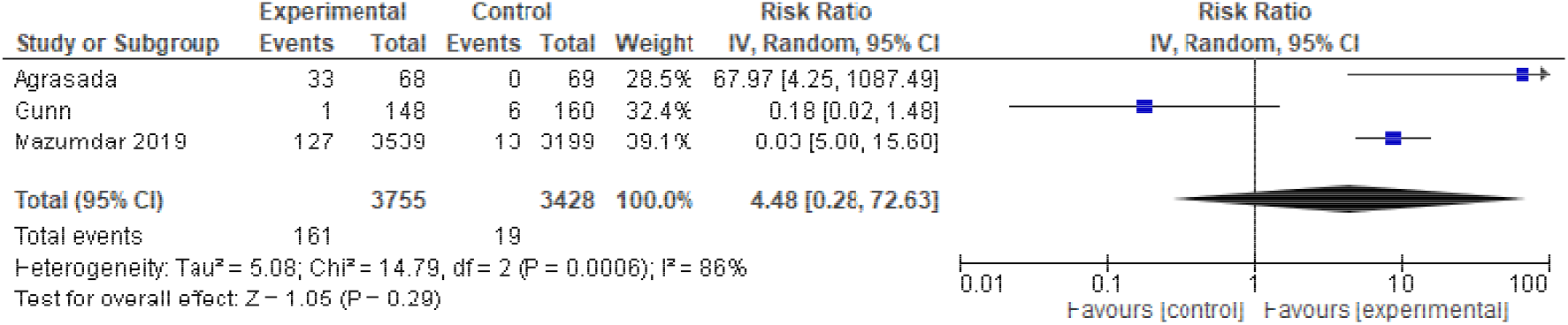
Exclusive breastfeeding.

Two RCTs (41, 42) reported no effect on cognitive development at 10-12 months corrected age and were included in meta-analysis (n=652, SMD 0.03, 95% CI −0.12 to 0.19).

**Figure 5:**
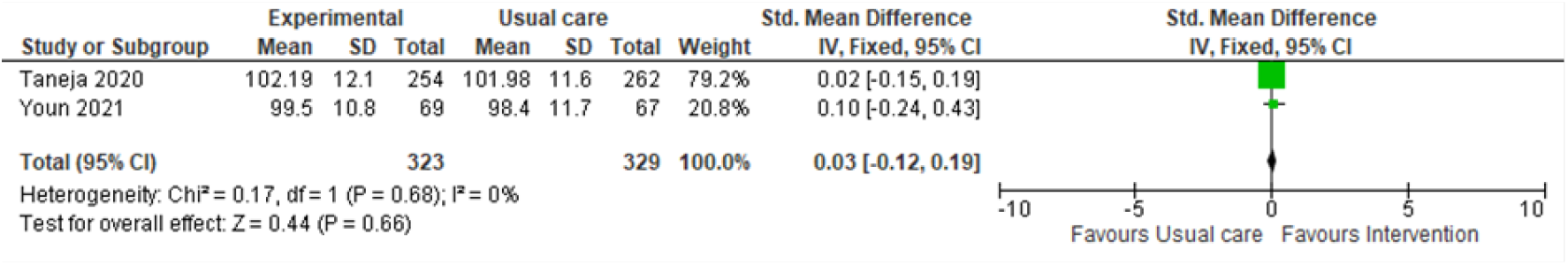
Infant cognitive development (measured by Bailey Scales of Infant Development III)

There was no evidence of effect on motor development at 10 months corrected age in one RCT (42) (n=136, MD 0.20, 95% CI 4.47 to 4.07) or infant temperament (21) (n=161, MD 0.70, 95% CI −0.6 to 1.46) at six months corrected age.

Five RCTs reported maternal outcomes including maternal depression (two studies), stress (two studies) and mother-infant attachment (one study).

A reduction in maternal depression was reported at 28 days post-birth in one RCT (40) (n=1147, RR 0.74, 95% CI 0.55 to 1.00). However, an RCT (42) reporting maternal depression at 6 months corrected age found no evidence of effect between groups (n=136, MD-1.30, 95% CI −3.50 to 0.90). There was no evidence of effect reported in one quasi-experimental study for maternal stress at 6 months (33) (n-56, MD −3.54 95% CI −16.21 to 9.13) or by one RCT at 12 months (37) corrected age (n=162, MD 1.69, 95% CI −3.18 to 6.56) or mother-infant attachment (42) (n=136, MD 1.20, 95% CI 2.79 to 0.39) at 6 months corrected age.

### Strengthened discharge preparedness interventions

Four studies were included in this category, with the only infant outcome reported being emergency hospital visits (one study).

Emergency hospital visits was the only infant outcome reported for this intervention type (Table 4). One observational study (43) reported that infants in the intervention group were less likely to attend for emergency care than those in the control group (n=173, RR 0.62, 95% CI 0.39-1.0) at two months follow up.

Three RCTsreported maternal outcomes including quality of life (one study) and stress (two studies) (Table 4). One reported an increase in maternal quality of life in the intervention group (n=56, MD 34.5, 95% CI 30.5 to 38.5) at four weeks follow up. Two RCTs reported no evidence of effect in maternal stress (44) (n= 26, MD −1.10, 95% CI −4.64 to 2.44) at one to two months or anxiety (46) (n=72, MD −2.30, 95% CI −5.49 to 0.89) at 3 months follow up.

### Digital communication interventions

A total of four RCTs were included in this category, with three studies reporting infant outcomes of exclusive breastfeeding (two studies) and emergency hospital visits (one study) (Table 5). Two studies reported no evidence of effect in exclusive breastfeeding (55, 56) at one to two months post-discharge (n=641, RR 1.02, 95% CI 0.89 to 1.16). One study reported emergency hospital visits (50) to be significantly reduced in the intervention group (n=89, median 0, range 0-7, compared with median 1 range 0-6) at 2 months post-discharge.

Two RCTs reported maternal-infant interaction as an outcome(49, 56). No evidence of effect was reported in either study at one month follow up (56) (n=129, MD −0.80, 95% CI −1.84 to 0.24) or 4 months of age (49) (n=85, MD −0.9, 95% CI −2.09 to 0.29).

### Peer support interventions

Two studies were included in this category, reporting one infant outcome of exclusive breastfeeding duration (one study) and one maternal outcome of anxiety (one study) (table 6).

One RCT (44)(n=69) reported no evidence of effect in duration of exclusive breastfeeding between groups, with the intervention group reporting a median of 3 months (range 0-14 months) and the control group a median of 4.3 months (range 0-13 months).

One observational study (45) (n=49) reported maternal anxiety to be lower in the intervention group (MD −7.20, 95% CI −12.56 to 1.84) at four months follow up.

## Discussion

This systematic review is the first, to our knowledge, to specifically address interventions to support parents in caring for preterm or LBW infants in the home. The early period following babies’ transfer into the community is a critical period for all babies; preterm and low birthweight babies are particularly vulnerable (57), thus parental support for caring for these babies is important (58).

This review identified multiple interventions aimed at offering such support, with evidence of significant effect size related to some outcomes. Studies reporting education and counselling interventions demonstrated improvements in infant growth, exclusive breastfeeding, infant cognitive development, and reduced maternal stress. Discharge preparedness interventions reported a significantly improved maternal quality of life. Home visiting interventions demonstrated a significant reduction in infant mortality, increase in immunisation uptake, and reduction in maternal depression, whilst peer-support interventions noted a significant reduction in maternal anxiety. These findings support the need for greater community support for women in caring for their babies (59), however the wide variation in interventions, the methodological weaknesses and inconsistencies in outcome measures highlights the need for further rigorous research prior to implementation of specific interventions.

Furthermore, although the findings are encouraging, none of the papers provided detailed process evaluation, therefore it is difficult to understand the mechanisms of effect of the interventions or the impact of contextual factors (60), making replicability challenging. Intervention fidelity and compliance were also generally ill-defined, with potential impact on outcomes (60). Inconsistencies in outcome measures used and the timing of assessments prevented meta-analysis of data for the majority of papers. This suggests that, for this area of investigation, development of core outcomes sets would be valuable, to enable consensus on a standardised minimal group of important outcomes (61).

Parental acceptability of the interventions received little attention in the included studies with only six papers reporting any aspects of acceptability (32, 43, 49–52). Where this was reported, the overall feedback from parents was positive. However, understanding acceptability is complex and dependent on several factors (62) and future studies should consider mixed methods of data collection, including in-depth qualitative explorations to capture the nuances of acceptability. For example, peer support was particularly well received (51, 52), but it is difficult to determine whether this was a result of the intervention per se or because of the additional social contact with other mothers. Moreover, none of the papers reported involving women or families in the development of interventions. Engagement and involvement of women, families and communities provides for more relevant and sustainable interventions (63).

The evidence presented in the included papers failed to provide confidence in the impact of the interventions on maternal psychological morbidity. Nevertheless, we know that outcomes such as stress, anxiety and depression are important to women when caring for their babies (7) suggesting further adequately powered, rigorous studies are required.

Although 37 studies were included in this review this was over a 22-year period with a diversity of interventions. Whilst a plethora of research has been conducted to prevent preterm or LBW infants (64), or to provide support and care in the hospital (65), support for parents to care for their preterm or LBW infants at home has been a neglected area. In many settings there is limited or no postnatal support for women after discharge regardless of infant gestation or weight (66). This is a particularly difficult time for parents, who have reported feeling vulnerable and inadequate in caring for their newborn when transitioning from a facility where support was available to being left to cope alone (6). It is conceivable that a well-developed programme of support, in the home, to enable families to care for preterm or LBW infants may improve health outcomes for the infant and family, providing parents with the confidence and skills to care for their infant. The evidence to support such a programme needs further investigation.

### Strengths and Limitations

This is the first review exploring multiple interventions for parental support of preterm and LBW babies in the home. The review highlights the gaps in the evidence and illuminates the urgent need for further research in this area.

This review was limited by methodological weakness of many studies. Most included studies had small sample sizes, with limited or no discussion of sample size calculation. Risk of bias was high, with unclear or inadequately described randomisation and allocation concealment in some studies.

Participant blinding was not possible in any of the included studies and assessor blinding was unclear in some studies. The overall certainty of evidence was rated as very low or low, with only three outcomes including evidence of moderate quality (exclusive breastfeeding, infant mortality, cognitive development), all related to home visiting interventions. Studies were mainly downgraded for risk of bias, indirectness and imprecision as a result of methodological issues, single site/setting and sample. The interventions themselves were disparate, with differences in intervention components (which, in some cases, were poorly described), methods of delivery, length and frequency of intervention. The outcome measures and timepoints at which these were measured also varied considerably between studies, even when the interventions were similar. Furthermore, no studies provided data from low-income settings.

## Conclusion

This review highlights the need for well-designed, effective support interventions, prioritised and developed with women, families and stakeholders. This will enable suitably tailored interventions which meet the needs of women and families, whilst improving infant outcomes. Components need to be carefully considered and evaluated in context, along with economic evaluation. Importantly, acceptability of interventions for women and families is vital in ensuring their success. Process evaluation and economic evaluation will also add valuable understanding to development and potential implementation of interventions. Furthermore, given the disparity of outcomes measured, development of a core outcome set for support interventions should be considered. The evidence does confirm the viewpoint that support interventions for parents to care for preterm or LBW infants in the home may improve outcomes. This is an area of research which requires further exploration, as outlined above, in order to further improve outcomes for preterm of LBW infants.

## Data Availability

All data produced in the present study are available upon reasonable request to the authors

## Notes

### Competing Interest Statement

Grant funding for research from the World Health Organisation through their organisation. All authors have completed the ICMJE uniform disclosure form at www.icmje.org/coi_disclosure.pdf. AP is employed by the World Health Organisation.

### Clinical Protocols

https://www.crd.york.ac.uk/prospero/display_record.php?ID=CRD42021275525

### Funding Statement

This study was funded by the World Health Organisation.

