## Supplementary material for "Interventions to support parents, families and caregivers in caring for preterm or low birth weight infants at home: a systematic review": Table 1

**Table 1: Summary of interventions**

| Author | Population | Intervention type | Intervention components and focus | Length of intervention | Delivered by | Outcome & measure |
| --- | --- | --- | --- | --- | --- | --- |
| Ahmed 2008 | P <37 | Education and counselling | Individual information giving and skills training in breastfeeding | 4 in NICU /1 home, birth to 3 months post-discharge | Researcher | Exclusive breastfeeding at 2-3 months |
| Dusing 2015, 2018 | P <37 | Education and counselling | Individual skills training in infant care and development | At least twice in NICU/ twice weekly at home from discharge, birth to 3 months CA | Trained and experienced NICU Physical therapist | Motor and cognitive development at 6 months; assessed with Bayley Scales of Infant Development (BSID) |
| Fan 2021 | P 28-32 | Education and counselling | Individual information giving and skills training in infant development | 1 training lecture and 3 workshops in the facility with parents performing EI in the home, for 30 days. | Researcher with rehabilitation background | Infant weight and length at 120 days follow-up |
| Finlayson 2020 | P <30 | Education and counselling | Individual skills training in infant care and development | 5 in NICU/ 5 home, birt to 3 months CA | Experienced Physiotherapist/ occupational therapists trained in intervention | Cognitive development at 4-6 months; assessed with BSID |
| Fotiou 2016 | P <37 | Education and counselling | Group information and skills training, with additional materials, in self-care | 5 sessions (90 minutes each) in NICU, parents practised independently for 3 months after discharge, birth to 3 months CA | Researcher | Maternal anxiety at 1-2 months; assessed with State Trait Anxiety Inventory (STAI) |
| Glazebrook 2007 | P <32 | Education and counselling | Individual information giving and skills training in sensitivity to infant | Weekly one hour sessions until discharge / optional to continue until 6 weeks following discharge | NICU nurse trained in intervention | Mother-infant interaction at 3 months; assessed with: Nursing Child Assessment Teaching Scale (NCATS). Maternal stress at 3 months follow-up; assessed with Parenting Stress Index (PSI) |
| Jaywant 2020 | P 28-36 | Education and counselling | Individual information giving and skills training in infant care and sensitivity to infant | Daily session in NICU, for 15 days | Therapist | Maternal stress at 15 days follow-up; assessed with parental stress scale (PSS). Maternal depression at 15 days; assessed with Edinburgh Postnatal Depression Scale (EPDS) |
| Kaaresen 2005 | LBW | Education and counselling | Individual information giving and skills training in self-care and sensitivity to infant | Daily one hour for 7 days pre-discharge / 4 home visits, 7 day pre- to 90 days post-discharge | NICU nurse trained to deliver intervention | Maternal stress at 12 months follow-up; assessed with PSI |
| Melnyk 2001 | P 26-36 & LBW | Education and counselling | Self-directed materials in infant care, development and sensitivity to infant | 4 phase programme, 2-4 days post-birth to 1 week post-discharge | Self-directed | Cognitive development at 4-6 months; assessed with BSID. Maternal anxiety at 1-2 months; assessed with State Trait Anxiety Inventory (STAI). Maternal depression at 6 months; assessed with Profile of Mood States |
| Milgrom 2016 | P <30 | Education and counselling | Individual information giving and skills training in self-care and sensitivity to infant | Weekly for 9 weeks in NICU / at home | Psychologist with experience of preterm populations | Infant temperament at 6 months; assessed by Short Temperament Scale |
| Moudi 2019 | P 23-37 | Education and counselling | Individual information giving and skills training, with materials, in care of infant | 4 sessions (60-90 mins each) in NICU. Researcher available for additional telephone support, 2-4 days post-birth to discharge | Researcher | Maternal anxiety at 1-2 months; assessed with State Trait Anxiety Inventory (STAI) |
| Newnham 2009 | P <37 | Education and counselling | Individual information giving and skills training in infant care, development and sensitivity to infant | 7 (30-60 min) sessions over final 2 weeks in NICU, 1 at home and 1 hospital visit, 2 weeks pre- to 3 months post-discharge | Researcher | Infant temperament at 6 months; assessed by Short Temperament Scale. Mother-infant interaction at 6 months; assessed with: Synchrony Scale. Maternal depression at 6 months; assessed with EPDS |
| Pinelli 2001 | VLBW | Education and counselling | Individual skills training, with materials, in breastfeeding | Weekly in NICU, ‘frequently’ at home after discharge, 72h following birth until 1 year or B/F discontinued | Independent lactation consultant | Duration of exclusive breastfeeding |
| Thakur 2012 | LBW | Education and counselling | Individual information giving in breastfeeding | 4 sessions (x2 per month from initiation of breastfeeding for 2 months) | Not stated | Infant weight and length at 60 days |
| White Traut 2013 | P 29-34 | Education and counselling | Individual information giving and skills training, with materials, in infant development and sensitivity to infant | Twice daily in NICU by mother or research nurse and continued at home by mother. Two facility sessions, two home visits and two telephone calls. Study entry (or 32 weeks) to 1 month post-discharge | Research nurse | Mother-infant interaction at 6 weeks; assessed with: Nursing Child Assessment Satellite Training–Feeding Scale (NCAST-Feeding) |
| Wu 2014 | P &  VLBW | Education and counselling | Individual information giving and skills training in infant care and development | 5 sessions NICU/ 8 home, within 7 days birth to 12 months CA | Nurse & physical therapist | Mother-infant interaction at 12 months follow-up; assessed with: Free-play procedure |
| Zelkowitz 2009 | VLBW | Education and counselling | Individual information giving, with materials, in self-care and sensitivity to infant | 5 (1 hour sessions NICU) from average 33 days post-birth / 1 home 2-4 weeks post- discharge. | Nurse, psychologist / graduate student trained to deliver intervention | Maternal anxiety at 6 months; assessed with STAI |
| Agrasada 2005 | LBW | Home visits | Individual information giving and skills training in breastfeeding. | 8 home visits, 3 days following birth until 5.5 months | Trained village volunteers | Exclusive breastfeeding at 6 months |
| Gardner 2003 | LBW | Home visits | Individual information giving, materials in sensitivity to infant. | Weekly 1 h home visits for 8 weeks | Community health workers | Bayley scales of infant development; at 10-12 months |
| Gunn 2000 | P <37 | Home visits | Individual breastfeeding support. | Daily visits for 7-10 days post-discharge. Telephone support available | Home care nurse specialists with neonatal experience | Number exclusively breastfeeding at 6 months |
| Ji & Shim 2020 | P <37 | Home visits | Individual and group information giving and skills training in care of and sensitivity to infant. | 1 or 2 home visits per month for 6 months, plus group support sessions | Experienced NICU nurse and a community visiting nurse | Maternal stress at 6 months; PSI |
| Koldewijn 2005, 2009 | P <32 | Home visits | Individual information giving, in care planning, infant development and sensitivity to infant. | 6-8 (1h) home visits as required. 1 week post-discharge to 6 months | Trained paediatric physical therapists | Infant temperament at 6 months; Infant behavioural assessment (IBA) |
| Mazumder 2019 | LBW | Home visits | Individual information giving and skills training in breastfeeding and KMC | 9 home visits (30-40mins) at 1-3, 5, 7, 10, 14, 21, 28 days | Intensively trained intervention worker (not HW) | Infant mortality up to 180 days  Number exclusively breastfeeding at 6 months |
| McKelvey 2021 | P & LBW | Home visits | Individual information giving in infant care | 2 home visits per month for 2 months, then 1 visit per month until 1 year | Registered nurse and licensed social worker | Infant mortality and hospitalisation until 12 months |
| Meijessen 2010a; 2020b | P or VLBW | Home visits | Individual information giving, in care planning, infant development and sensitivity to infant. | 6-8 (1h) home visits as required until 12 months CA | Experienced trained paediatric physical therapist | Maternal stress at 12 months; PSI |
| Sinha 2021 | LBW | Home visits | Individual information giving and skills training in breastfeeding and KMC | 9 home visits (30-40mins) at 1-3, 5, 7, 10, 14, 21, 28 days | Intensively trained intervention worker (not HW) | Maternal depression at 28 days; Patient health questionnaire 9. |
| Taneja 2020 | LBW | Home visits | Individual information giving and skills training in breastfeeding and KMC | 9 home visits (30-40mins) at 1-3, 5, 7, 10, 14, 21, 28 days | Intensively trained intervention worker (not HW) | Cognitive development. Bayley scales of infant development; at 10-12 months |
| Youn 2021 | P <30 or VLBW | Home visits | Individual information giving and group skills training in infant care, development and sensitivity to infant. | 4 home visits (Discharge until 2 months CA) 12 x 90 min group sessions with physiotherapist | Specialist nurse and physiotherapist (infant neurodevelopment) | Cognitive and motor development at 10 months; . Bayley scales of infant development  Mother/infant attachment at 6 months; Mother-Child Attachment (MCA) scale |
| Ingram 2016 | P 27-<34 | Discharge preparedness | Planning for early discharge. Care of infant. | As required, for 5 weeks, between 27-33 gw | NICU staff | Emergency hospital visits; number until 2 months |
| Lee 2019 | P <32 | Discharge preparedness | Planning for discharge. Care of infant. | 3 sessions and 1 FU tele call. 34 gw to 72h post-discharge | Neonatal nurse consultant | Maternal stress 1-2 months; assessed with Perceived Stress Scale (C-PSS) |
| Neyestani 2017 | P 30-35 | Discharge preparedness | Planning for discharge. Care of infant.  Materials. Included parent-directed contact following discharge if required. | 4 (35-40 min) sessions plus 4 telephone contacts (x1 per week, 5-10 mins) after discharge. 48h from birth until 4wks post-discharge. | Nurse | Maternal quality of life at 4 weeks; assessed with WHOQOL-BREF |
| Ortenstrand 2001 | P <37 | Discharge preparedness | Planning for discharge and early discharge. Care of infant | Care planning session and domiciliary care visits (number unclear). Pre-discharge until domiciliary care completed. | Project nurse (neonatal trained) | Maternal anxiety at 3 months; assessed with STAI |
| Ericson 2018 | P <37 | Digital communication | Individual support/communication in breastfeeding | Daily telephone call from discharge for 14 days | Breastfeeding support team (NICU staff trained for 2 days) | Exclusive breastfeeding at 1-2 months |
| Hagi-Pederson 2020 | P <37 | Digital communication | Individual support/communication in breastfeeding | 2-3 consultations per week by video following introduction in NICU, until discharge programme completion | Nurse trained in early in-home care and trained in smartphone application, video consultations | Exclusive breastfeeding at 1-2 months. Maternal-infant interaction at 1 month; assessed with: Mother and Baby Interaction Scale (MABISC) |
| Luu 2017 | P <30 | Digital communication | Individual support/communication, with materials, in infant care | 3x in-person workshop and 4 web based modules (commenced in NICU), until 12 months | Certified occupational therapist trained in | Maternal-infant interaction at 4 months; assessed with: Parental Cognitions and Conduct Toward the Infant Scale |
| Robinson 2016 | P 27-37 | Digital communication | Individual support/communication in infant care | 3x Skype calls per week with messaging option, to discharge from home health care | developmental care. | Emergency hospital visits up to 2 months post-discharge |
| Neila-Vilen  2016 | P <35 | Peer support | Individual support and materials in breastfeeding | As required (facility / home), to 12 months | Volunteers with experience of B/F preterm infants (no training given).  Midwife available to answer B/F questions. | Duration of exclusive breastfeeding |
| Preyde 2003 | P <30 | Peer support | Individual support and support group | Average 9 contacts (facility/ home), within week of birth to 16 weeks | Trained mothers with experience of caring for preterm infants | Maternal anxiety (STAI) at 4 months |
