## Supplementary material for "Interventions to support parents, families and caregivers in caring for preterm or low birth weight infants at home: a systematic review": Table 2

**Table 2: Education and counselling interventions**

| Outcomes | **Mean (SD) or No. (%)** | | **Relative effect (95% CI)** | **№ of participants (studies)** | **Certainty of the evidence (GRADE)** |
| --- | --- | --- | --- | --- | --- |
|  | **usual care** | Facility-initiated education and counselling |  |  |  |
| **Infant length  at 60 days**  follow-up (cm); Mean (SD)  Thakur 2012 | 48.7 (1.6) | 50.2 (1.3) | MD 1.5 cm higher (1.1 cm higher to 1.9 cm higher) | 184  (1 RCT) | ⨁◯◯◯ Very low ^a, b, c, e^ |
| **Infant length  at 120 days** follow-up (cm); Mean (SD)  Fan 2021 | 58.6 (2.6) | 59.8 (2.8) | MD 1.2 cm higher  (0.2 cm higher to 2.6 cm higher) | 57  (1 RCT) | ⨁◯◯◯ Very low ^e,h,i^ |
| **Infant weight  at 60 days**  follow-up (g); Mean (SD) Thakur 2012 | 3315 (300) | 3620 (229) | MD 305 g higher (228 g higher to 382 g higher) | 184  (1 RCT) | ⨁◯◯◯ Very low ^a,b,c,e^ |
| **Infant weight at 120 days** follow-up (kg); Mean (SD)  Fan 2021 | 5.24 (0.95) | 5.56 (0.92) | MD 410 g higher (406 g higher to 414 g higher) | 57  (1 RCT) | ⨁◯◯◯ Very low ^e ,h, i^ |
| **Exclusive breastfeeding** at 2-3 months; n (%)  Ahmed 2008  Thakur 2012 | 38/122 (31.1%) | 67/122 (54.9%) | RR 1.71 (1.26 to 2.31) | 244 (2 RCTs) | ⨁◯◯◯ Very low ^a,b,c^ |
| **Duration of exclusive breastfeeding**; Mean (SD)  Pinelli 2001 | 24.2 weeks (21.6 weeks) | 26.2 weeks (21.6 weeks) | MD 2.0 weeks higher (5.48 weeks lower to 9.48 weeks higher) | 128  (1 RCT) | ⨁◯◯◯ Very low ^b, d, e^ |
| **Motor development at 6 months**; assessed with Bayley Scales of Infant Development (BSID); Mean (SD)  Dusing 2015 | NA | NA | SMD 0.38 higher (1.15 lower to 1.91 higher) | 7 (1 RCT) | ⨁◯◯◯ Very low ^a,d,e,f^ |
| **Cognitive development at 4-6 months**; assessed with BSID (II or III) Mean (SD)  Dusing 2015  Finlayson 2020  Melnyk 2001 | NA | NA | SMD 0.67 higher (0.16 higher to 1.17 higher) | 65 (3 RCTs) | ⨁◯◯◯ Very low ^a, d, g^ |
| **Infant temperament at 6 months**; assessed by Short Temperament Scale  Newnham 2009, Milgrom 2013 | NA | NA | MD 0.54 higher  (0.06 lower to 1.02 higher) | 155  (2 RCTs) | ⨁⨁◯◯ Low^a, d^ |

| Mother-infant interaction at 6 weeks; assessed with: Nursing Child Assessment Satellite Training–Feeding Scale (NCAST-Feeding); Mean (SD)  White-Traut 2013 | 62.5 (7.0), | 64.3 (5.2) | MD 1.80 higher (0.21 higher to 3.81 higher) | 142  (1 RCT) | ⨁⨁◯◯ Low ^d,e^ |
| --- | --- | --- | --- | --- | --- |
| Mother-infant interaction at 3 months; assessed with: Nursing Child Assessment Teaching Scale (NCATS); Mean (SD)  Glazebrook 2007 | 37.4 (4.9) | 36.6 (5.1) | MD 0.80 (2.20 higher to 0.60 higher) | 196  (1 RCT) | ⨁⨁◯◯ Low ^d,e^ |
| Mother-infant interaction at 6 months; assessed with: Synchrony Scale; Mean (SD)  Newnham 2009 | 0.24, (0.13) | 0.45 (0.08) | MD 21.0 (0.11 higher to 0.67 higher) | 63  (1 RCT) | ⨁◯◯◯ Very Low ^d,e^ |
| Mother-infant interaction at 12 months follow-up; assessed with: Free-play procedure; Mean (SD)  Wu 2014   - High quality maternal behaviour        - Engaged infant behaviour      - Synchronous didactic behaviour | 0.41 (0.27))      0.85 (0.19)      0.35 (0.25) | CHIB: 0.51 (0.28) HBIP: 0.46 (0.29)      CBIP 0.86 (0.13) HBIP 0.89 (0.15)    CBIP 0.44 (0.24) HBIP 0.42 (0.28) | MD 0.10 higher (0.01 lower to 0.21 higher)  MD 0.01 lower (0.06 lower to 0.08 higher)  MD 0.09 higher (0.01 lower to 0.19 higher). | (93)  1 RCT | ⨁⨁◯◯ Low ^c,e^ |
| Maternal stress at 15 days follow-up; assessed with parental stress scale (PSS)  Jaywant 2020 | NA | NA | MD 1.12 lower (14.32 lower to 12.08 higher) | 52  (1 RCT) | ⨁◯◯◯ Very Low ^b,d^ |
| Maternal stress at 3 months follow-up; assessed with Parenting Stress Index (PSI) Glazebrook 2007 | NA | NA | MD 4.80 higher (0.56 lower to 10.16 higher) | 199  (1 RCT) | ⨁⨁◯◯ Low ^b,d^ |
| Maternal stress at 12 months follow-up; assessed with PSI Kaaresen 2005 | NA | NA | MD 13.70 lower (25.5 lower to 1.89 lower) | 130  (1 RCT) | ⨁⨁◯◯ Low ^d,e^ |
| Maternal anxiety at 1-2 months; assessed with State Trait Anxiety Inventory (STAI)  Moudi 2019  Melnyk 2001 | NA | NA | SMD 1.89 lower (5.31 lower to 1.53 higher) | 177  (2 RCTs) | ⨁⨁◯◯ Low ^j,k^ |
| Maternal anxiety at 1-2 months; assessed with State Trait Anxiety Inventory (STAI)  Fotiou 2016 | NA | NA | MD 1.50 lower  (9.35 lower to 6.35 higher) | 29  (RCT) | ⨁◯◯◯ Very low ^b,d^ |
| Maternal anxiety at 6 months; assessed with STAI  Zelkowitz 2009 | NA | NA | MD 1.20 lower (4.18 lower to 1.78 higher) | 42  (1 RCT) | ⨁⨁◯◯ Low ^d,e^ |
| Maternal depression at 15 days; assessed with Edinburgh Postnatal Depression Scale (EPDS)  Jaywant 2020 | NA | NA | MD 0.64 lower (3.53 lower to 2.25 higher) | 52  (1 RCT) | ⨁◯◯◯ Very Low ^a,b,c,e^ |
| Maternal depression at 6 months; assessed with Profile of Mood States (POMS)/EPDS)  Melnyk 2001  Newnham 2009 | NA | NA | SMD 0.25 lower (0.65 lower to 0.16 higher) | 94  (2 RCTs) | ⨁◯◯◯ Very Low ^b,d,h^ |

a. Randomisation and allocation concealment unclear or not described. b. Blinding of assessors unclear c. Lower-middle income setting. d. Single high-income setting. e. Single study. f. Attrition >10%. g. Small sample size. h. Allocation concealment unclear. i. Upper middle-income setting j. Randomisation and allocation concealment unclear in one study. k. Heterogeneity. Wide variation in CI between studies.
