## Supplementary material for "Interventions to support parents, families and caregivers in caring for preterm or low birth weight infants at home: a systematic review": Table 3

**Table 3: Home visiting interventions**

| Outcomes | **Absolute effects** | | Relative effect (95% CI) | № of participants (studies) | Certainty of the evidence (GRADE) |
| --- | --- | --- | --- | --- | --- |
|  | **Usual care** | **Home visits** |  |  |  |
| Mortality until 180 days Mazumdar 2019 | 47 per 1,000 | 35 per 1,000 (28 to 44) | RR 0.71 (0.57 to 0.89) | 7479 (1 RCT) | ⨁⨁⨁◯ Moderate^g,e^ |
| Mortality until 12 months; McKelvey 2021 | 14 per 1,000 | 2 per 1000  (0 to 17) | RR 0.14  (0.02 to 1.16) | 970  (1 observational study) | ⨁◯◯◯ Very low^c,d,e^ |
| Exclusive breastfeeding at 6 months  Agrasada 2005  Gunn 2000  Mazumder 2019 | 19/3428 (0.55%) | 161/3755 (4.3%) | RR 4.48 (0.28 to 72.9) | 7221 (3 RCTs) | ⨁⨁⨁◯ Moderate^a,b^ |
| Immunization visits in the first year of life; Mean (SD)  McKelvey 2021 | 2.53 (0.11) | 3.74 (0.09) | MD 1.21 higher  (0.93 higher to 1.49 higher) | 970  (1 observational study) | ⨁◯◯◯ Very low^,c,d,e^ |
| Hospitalization until 12 months FU; mean (SD)  McKelvey 2021 | 0.25 (0.88) | 0.59 (1.76) | MD 0.34 higher  (0.16 higher to 0.52 higher) | 970  (1 observational study) | ⨁◯◯◯ Very low^c,d,e^ |
| Cognitive development at 10-12 months; assessed by BSID  Taneja 2020  Youn 2021 | NA | NA | SMD 0.03 higher (0.12 lower to 0.19 higher) | 652  (2 RCTs) | ⨁⨁⨁◯  Moderate^a,b^ |
| Motor development  at 10 months; assessed by BSID  Youn 2021 | NA | NA | MD 0.20 lower (4.47 lower to 4.07 higher) | 136 (1 RCT) | ⨁⨁◯◯ Low^e,f^ |
| Infant temperament at 6 months; assessed by Infant behavioural assessment (IBA)  Koldewijn 2009 | NA | NA | MD 0.70 higher  (0.6 lower to 1.46 higher) | 161  (1 RCT) | ⨁⨁◯◯ Low^f,e^ |
| Mother-infant attachment (MCA); at 6 months Mean (SD)  Youn 2021 | 101.3 (5.1) | 100.1 (4.3) | MD 1.20 higher  (2.79 higher to 0.39 higher) | 136  (1 RCT) | ⨁⨁◯◯ Low^e,f^ |
| Maternal stress at 6 months; assessed with PSI. Mean (SD)  Ji & Shim 2020 | 80.81 (24.11) | 77.27 (24.24) | MD 3.54 lower  (16.21 lower to 9.13 higher) | 56  (1 observational study) | ⨁◯◯◯ Very low^e,f,h^ |
| Maternal stress at 12 months; assessed with PSI  Meijssen 2010b | N/A | N/A | MD 1.69 higher (3.18 lower to 6.56 higher) | 162  (1 RCT) | ⨁⨁◯◯ Low^e,f^ |
| Maternal depression at 28 days (PT and LBW); assessed by Patient Health Questionnaire – 9  Sinha 2021 | N/A | N/A | RR 0.74 (0.55 to 1.00) | 1147  (1 RCT) | ⨁⨁◯◯ Low^e,g^ |
| Maternal depression at 6 months; assessed  Youn 2021 | N/A | N/A | MD 1.30 lower (3.50 lower to 0.90 higher) | 136  (1 RCT) | ⨁⨁◯◯ Low^e,f^ |

a. Randomisation and allocation concealment not described, in one study. b. Binding of assessors not clear, in one study. c. Non-randomised, quasi-experimental study. Matched control group. d. Single high-income setting, six sites. e. Single study. f. High income setting. g. Lower middle-income setting h. Non-randomised, non-equivalent control group
