## Supplementary material for "Interventions to support parents, families and caregivers in caring for preterm or low birth weight infants at home: a systematic review": Table 4

**Table 4: Strengthened discharge preparedness interventions**

| Outcomes | **Effects** (95% CI) | | Relative effect (95% CI) | № of participants (studies) | Certainty of the evidence (GRADE) |
| --- | --- | --- | --- | --- | --- |
|  | **usual care** | **Strengthened discharge preparedness** |  |  |  |
| Emergency hospital visits until 2 months follow-up (Number of visits)  Ingram 2016 | 31/85 | 20/88 | **RR 0.62**  (CI 0.39 to 1.0) | 173  (1 observational study) | ⨁◯◯◯ Very low^a,b^ |
| Maternal stress at 1-2 months; assessed with Perceived Stress Scale (C-PSS)  Lee 2019 | NA | NA | MD **1.10 lower** (4.64 lower to 2.44 higher) | 26 (1 RCT) | ⨁⨁◯◯ Low^a,b^ |
| Parent anxiety -at 3 months; assessed with STAI  Ortenstrand 2001 | NA | NA | MD **2.30 lower** (5.49 lower to 0.89 higher) | 72 (1 RCT) | ⨁◯◯◯ Very low ^b, c,d^ |
| Maternal quality of life at 4 weeks follow-up; assessed with WHOQOL-BREF  Neyestani 2017 | 56.4 (4.1) | 90.9 (9.9) | MD 34.5 higher (30.5 higher to 38.5 higher) | 56  (1 RCT) | ⨁◯◯◯ Very low^d,,e, f, g^ |

a. Before and after study. Control group recruited prior to introduction of intervention. b. Single high-income setting. c. Randomisation and allocation concealment not adequately described. d. Blinding of assessors not clear e. Single lower middle-income country. f. Not all outcomes reported.
