## Supplementary material for "Interventions to support parents, families and caregivers in caring for preterm or low birth weight infants at home: a systematic review": Table 5

**Table 5: Digital communication interventions**

| Outcomes | **Effects** | | Relative effect (95% CI) | № of participants (studies) | Certainty of the evidence (GRADE) |
| --- | --- | --- | --- | --- | --- |
|  | **usual care** | **Digital communication** |  |  |  |
| Exclusive breastfeeding at 1-2 months  Ericson 2018  Hagi-Pederson 2020 | 207/361 | 185/327 | RR 1.02 (0.89 to 1.16) | 641 (2 RCTs) | ⨁◯◯◯ Very low^a,b,c^ |
| Emergency hospital visits up to 2 months post-discharge. Median (range)  Robinson 2016 | Median 1  (0-6) | Median 0  (0-7) |  | 89  (1 RCT) | ⨁◯◯◯ Very low^a,c,d,e^ |
| Maternal-infant interaction at 1 month follow-up; assessed with: Mother and Baby Interaction Scale (MABISC); Total score, Mean (SD**)**  Hagi-Pederson 2020 | 11.3 (3.4) | 10.5 (3.1) | MD 0.80 lower  (1.84 lower to 0.24 higher) | 129  (1 RCT) | ⨁◯◯◯ Very low^a,c,f^ |
| Maternal-infant interaction at 4 month follow-up; assessed with: Parental Cognitions and Conduct Toward the Infant Scale (PACOTIS); Median (IQR)  Luu 2017 | 9.0 (7.2-10.0) | 8.1 (7.0-9.8) | MD -0.9 lower (-2.09 lower to –0.29 higher)  P 0.59). | 85  (1 RCT) | ⨁◯◯◯ Very low^c, g^ |

a. Blinding of assessors not clear in one study b. Small sample size in one study c. High-income setting d. Randomisation and allocation concealment unclear/not described e. Single study f High attrition >10% g. Non-randomised, historical comparison group
