## Supplementary material for "Interventions to support parents, families and caregivers in caring for preterm or low birth weight infants at home: a systematic review": Table 6

**Table 6: Peer support interventions**

| Outcomes | **Effects** | | Relative effect (95% CI) | № of participants (studies) | Certainty of the evidence (GRADE) |
| --- | --- | --- | --- | --- | --- |
|  | **usual care** | **Facility-initiated peer support** |  |  |  |
| Duration of exclusive breastfeeding; Median (range)  Neila-Vilen 2016 | 4.3 months (range 0-13) in the control group. | 3 months (range 0-14) in the intervention group | - | 69  (1 RCT) | ⨁◯◯◯ Very low^a,b^ |
| Maternal anxiety (STAI) at 4 months;  Preyde 2003 | - | MD 7.20 lower (12.56 lower to 1.84 lower) | - | 49 (1 observational study) | ⨁◯◯◯ Very low^a,b,c^ |

a. Unclear if outcome assessors blinded. b. Single high-income setting c. Non-randomised cohort study with control group, >10% attrition
