## Supplementary file 1 for "Interventions to support parents, families and caregivers in caring for preterm or low birth weight infants at home: a systematic review"

**Interventions to support parents, families and carers in caring for premature or low birth weight (LBW) infants in the home**

**MEDLINE**

Search #1

preterm (kw) OR prematur* (kw) OR low N3 birthweight (kw) OR low N3 birth weight (kw) OR “high risk infant” (kw) OR Premature Birth (MeSH) OR Infant, Premature (MeSH) OR Infant, Extremely Premature (MeSH) OR Infant, Low Birth Weight (MeSH) OR Infant, Very Low Birth Weight (MeSH) OR Infant, Extremely Low Birth Weight (MeSH)

**AND**

Search #2

“post-discharge” (kw) OR postdischarge (kw) OR “patient discharge” (kw) OR “hospital discharge” (kw) OR facility discharge (kw) OR home (kw) OR household (kw) OR transition (kw) OR “self-efficacy” (kw) OR empower* (kw) OR Patient Discharge (MeSH) OR Transitional Care (MeSH) OR Self Efficacy (MeSH) OR Empowerment (MeSH)

**AND**

Search #3

mother (kw) OR father (kw) OR parent* (kw) OR caregiver (kw) OR “maternal care” (kw) OR famil* (kw) OR carer (kw) OR OR Caregivers (MeSH) OR Mothers (MeSH) OR Fathers (MeSH) OR Family (MeSH) OR Parents (MeSH) OR Parenting (MeSH) OR Mother-Child Relations (MeSH)

**AND**

Search #4

Support (kw) OR help (kw) OR Assistance OR intervention (kw) OR education (kw) OR Model OR partnership (kw)

Kw = keyword
MeSH = Medline subject heading
